# AI-HOPE: An AI-Driven conversational agent for enhanced clinical and genomic data integration in precision medicine research

**DOI:** 10.1101/2024.11.27.24318113

**Authors:** Ei-Wen Yang, Enrique Velazquez-Villarreal

## Abstract

Introduction: The increasing complexity of clinical cancer research necessitates the development of automated tools capable of integrating clinical and genomic data while accelerating discovery efforts. Artificial Intelligence agent for High-Optimization and Precision mEdicine (AI-HOPE) is introduced as an innovative conversational AI platform powered by Large Language Models (LLMs), designed to empower domain experts to perform integrative data analyses through natural language input, eliminating the need for programming expertise. AI-HOPE offers robust analytical capabilities, enabling the generation of actionable insights in clinical and translational research. Methods: AI-HOPE interprets user instructions in natural language and translates them into executable code to analyze locally stored data. It facilitates subset comparisons for clinical prevalence and survival analysis, generating statistical outputs such as odds ratios, Kaplan-Meier survival curves, and hazard ratios. Its capabilities were demonstrated through two case-control studies using The Cancer Genome Atlas (TCGA): (1) analyzing TP53 mutation enrichment in early-stage versus late-stage colorectal cancer (CRC) patients, and (2) comparing progression-free survival among FOLFOX-treated patients with or without RAS mutations. Results: In the first study, AI-HOPE identified a significant enrichment of TP53 mutations in late-stage (III/IV) CRC compared to early-stage (I/II) cases. In the second study, AI-HOPE revealed a significant association between KRAS mutations and poorer progression-free survival in FOLFOX-treated patients. These findings align with established literature, demonstrating AI-HOPE’s capability to independently uncover meaningful insights without prior user assumptions. Conclusions: AI-HOPE represents a transformative advancement in precision medicine research, offering a scalable, user-friendly framework for integrating clinical and genomic data. Its versatility extends beyond cancer research, supporting applications across diverse biomedical fields. Future enhancements, such as real-time data integration and multi-omics capabilities, will further solidify its role as a pivotal resource for advancing translational research and improving patient outcomes. AI-HOPE bridges the gap between data complexity and research needs, accelerating discoveries in precision medicine research.

## Introduction

Precision medicine is revolutionizing healthcare by tailoring treatments to individual genetic, environmental, and lifestyle factors (Collins et al., 2015; Jameson et al., 2015). At the core of this approach lies the integration of clinical and genomic data, providing a comprehensive understanding of disease mechanisms and therapeutic responses.

However, the vast volume and complexity of such data present significant challenges for clinical research (Tatonetti et al., 2019; Ashley et al., 2015). Extracting meaningful insights requires advanced bioinformatic analyses to interpret genomic data alongside clinical metadata (e.g., age, gender, tumor stage) and outcomes (e.g., overall survival, progression-free survival) (Ritchie et al., 2015; Katsanis et al., 2013). Unfortunately, traditional bioinformatics tools often require programming expertise, posing a barrier for many clinical researchers due to time constraints and technical demands.

To address these challenges, automated tools capable of end-to-end bioinformatics analyses are urgently needed to streamline clinical research workflows. Recent advances in Large Language Models (LLMs) have revolutionized artificial intelligence applications in biology, enabling transformative capabilities such as disease diagnosis (Wang et al., 2023) and drug discovery (Flam-Shepherd et al., 2022). A defining feature of LLMs is their conversational interface, which facilitates direct user interaction for complex tasks, moving beyond traditional GUI-based platforms to fully automated systems (Zhou et al., 2024). These systems, such as BIA and CellAgent, have demonstrated efficacy in automating single-cell RNA sequencing data processing (Xin et al., 2024; Xiao et al., 2024), while tools like AutoBA offer conventional multi-omics analysis capabilities (Zhou et al., 2024).

Despite these advances, existing LLM-based tools are limited in their ability to integrate clinical metadata with genomic data and treatment outcomes. Moreover, the flexibility to handle complex, user-defined data subsets is lacking, yet such capabilities are essential for clinical research. For example, identifying biomarkers, understanding disease progression, and evaluating treatment efficacy require the ability to compare groups based on clinical and molecular characteristics.

To address these limitations, we present the Artificial Intelligence agent for High-Optimization and Precision mEdicine (AI-HOPE), an innovative conversational AI platform designed to bridge these gaps. AI-HOPE leverages LLM technology to empower researchers with user-friendly tools for integrative data analysis, eliminating the need for programming expertise while enabling robust and flexible analytical workflows. This platform aims to advance translational research and accelerate discoveries in precision medicine.

## Methods

### AI-HOPE agent for precision medicine in clinical research

This workflow starts with the selection of case and control samples through natural language queries and then conduct key bioinformatics analyses, odds ratio tests and survival analysis. The process begins with the precise identification of clinically relevant case and control samples to establish a robust study design. Next, AI-HOPE agents execute the core analyses with minimal manual intervention. Finally, LLM agents synthesize the statistical results into detailed reports, providing clear insights for interpretation (Fig. 1).

**Figure 1.**
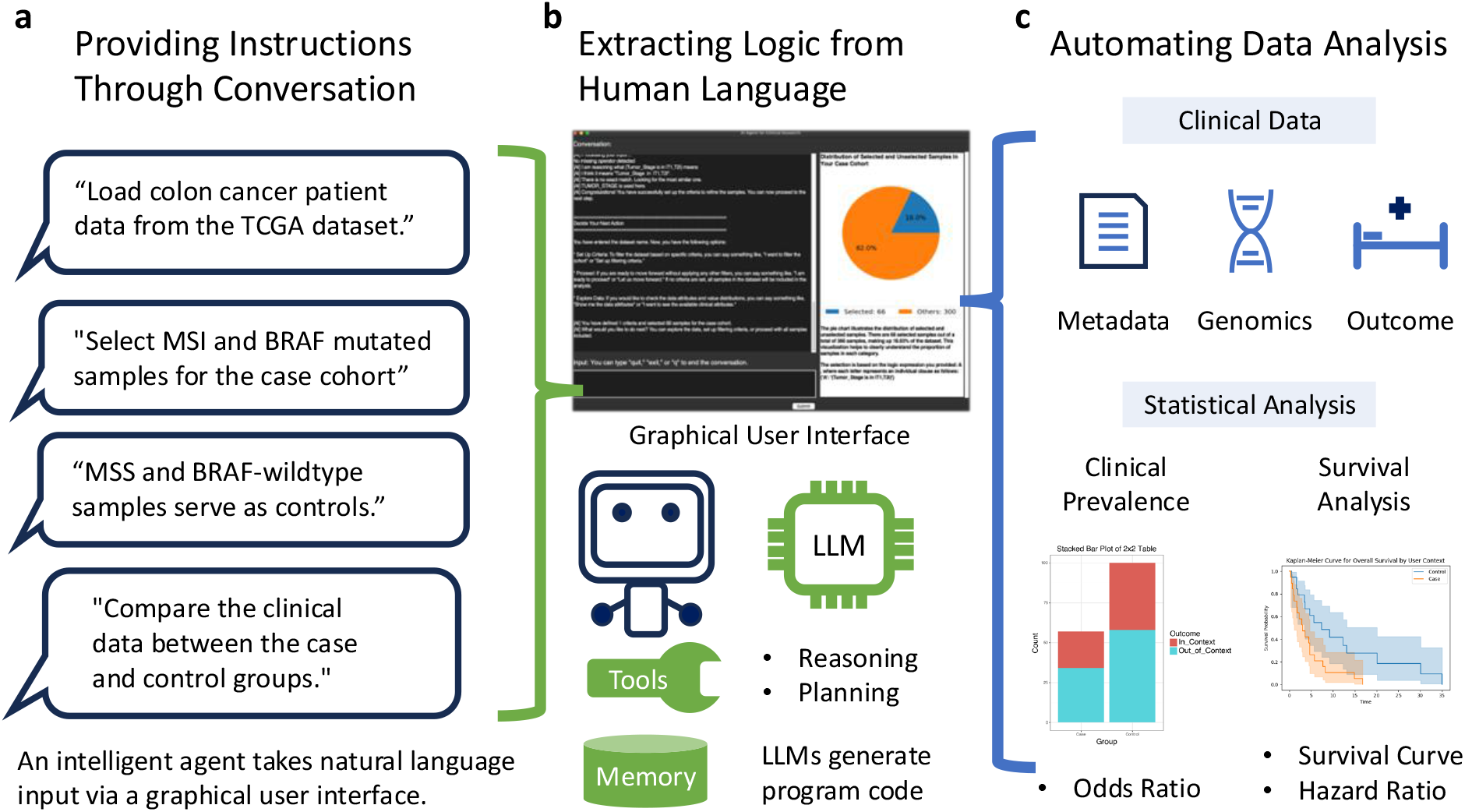
Overview of Artificial Intelligence agent for High-Optimization and Precision mEdicine (AI-HOPE) Workflow. This figure illustrates the workflow of the AI-HOPE system for clinical and genomic data analysis through a conversational interface. (a) Providing Instructions Through Conversation: The AI agent receives natural language instructions from the user, such as loading specific colon cancer patient data, selecting sample cohorts based on genomic features (e.g., MSI, BRAF mutations), and comparing clinical data between case and control groups. (b) Extracting Logic from Human Language: The agent processes the user’s inputs via a graphical user interface (GUI) that leverages a large language model (LLM). The LLM generates the necessary program code to execute tasks, utilizing memory and reasoning tools to ensure accurate data extraction and manipulation. (c) Automating Data Analysis: The system automates bioinformatics tasks, integrating clinical, genomic, and outcome data. Statistical analyses, such as clinical prevalence and survival analysis (e.g., odds ratio, survival curves, hazard ratio), are performed, with results displayed for interpretation and further research.

### Data Preparation

All datasets are installed separately in a designated folder. Each dataset contains a main data table in a tab-delimited text file, a README file explaining the data attributes to users, and an index file specifying the file names and key data attributes, such as Overall Survival (OS) and Progression-Free Survival (PFS). To demonstrate the value of AI-HOPE agents in clinical research, we integrate TCGA data from UCSC Xena (https://xena.ucsc.edu) and cBioPortal (accessed on 8 August 2024), covering 33 cancer types. Users can conveniently download the datasets using the provided URL.

### Load and Subset Data Using Natural Language

The workflow begins by prompting users to provide the name of the dataset for either case or control samples. Once the dataset name is entered, the agent facilitates sample selection and analysis setup. Users can refine their sample selection by providing criteria as natural language statements, which our pretrained LLM model converts into precise arithmetic expressions. Each statement must include a data attribute, a comparison operator, and a value. For instance, in the statement “Age is greater than 30,” “Age” represents the data attribute, “is greater than” is the comparison operator, and 30 is the value. Supported comparison operators include equality (e.g., “is,” “is not”), inequality (e.g., “greater than,” “less than”), ranges (e.g., “from [Start Value] to [End Value]”), and set inclusion or exclusion (e.g., “is in,” “is not in”). For example, the expression “Disease stage is in {stage I, stage II, stage III}” specifies a set of valid values for inclusion. For users unfamiliar with the data, the agent can assist in exploring data attributes and value distributions to guide the selection process. By following these guidelines, users can ensure complete accuracy in the model’s conversion of natural language statements into arithmetic expressions.

When defining multiple criteria, each logical clause should be enclosed in parentheses for clarity. Clauses can be connected using logical operators such as “and” or “or,” enabling the creation of complex expressions. For example, (Age is greater than 30) and (Gender is male) or (Diagnosis is cancer) combines multiple conditions. The input expression is evaluated in the postfix order of the clauses based on their position in the expression. Nested parentheses can be used to override the default evaluation order, ensuring specific priorities (see Methods in supplementary materials). This intuitive approach simplifies dataset subsetting, allowing researchers to interact seamlessly with the data using natural language.

### Autonomous Data analysis

The AI-HOPE agents offer two core analysis options: the Odds Ratio Test and survival analysis, both implemented using standard Python libraries for statistical analysis. The Odds Ratio Test calculates the odds ratio based on user-defined contexts, enabling researchers to assess the strength of association between case and control groups.

Users define specific contexts for analysis, following a process similar to defining case and control groups. The test evaluates the number of samples within and outside the defined context in each group using a 2×2 contingency table analyzed with a Chi-Square test.

The survival analysis utilizes user-provided case-control stratification to generate Kaplan-Meier (KM) plots for survival data, illustrating survival probabilities across user-defined groups. Statistical significance between survival curves is determined using the log-rank test, while Cox regression calculates hazard ratios (HRs) to assess the impact of case-control groups and additional variables on survival outcomes. Users can perform univariate analysis focused on case-control groups or extend to multivariate analysis by incorporating additional variables to examine combined effects on survival.

## Results

The AI-HOPE demonstrated its ability to interpret natural language user instructions, translating them into executable code to analyze locally stored clinical and genomic data. Through its conversational interface, AI-HOPE effectively classified patient samples into case and control cohorts based on user-defined criteria. These criteria allowed flexible stratification of patient groups using attributes such as genetic mutations, disease stages, and treatment responses. AI-HOPE autonomously conducted clinical prevalence and survival analyses for these cohorts, providing detailed statistical outcomes, including odds ratios, Kaplan-Meier survival curves, and hazard ratios. Beyond numerical outputs, AI-HOPE employed its LLM to interpret the results, offering actionable insights to streamline the research process.

In one use case, AI-HOPE analyzed colorectal cancer (CRC) data from The Cancer Genome Atlas (TCGA) to investigate tumor evolution. The agent performed a case-control study comparing TP53 mutations in early-stage (I/II) versus late-stage (III/IV) CRC patients. The system identified that 15.6% of patients were in the early-stage cohort and 32.2% in the late-stage cohort, revealing a significant enrichment of TP53 mutations in late-stage CRC. This finding aligns closely with existing literature, such as the observations of Lacopetta et al. (2006), further validating the reliability of AI-HOPE’s analytical capabilities (Fig. 2).

**Figure 2:**
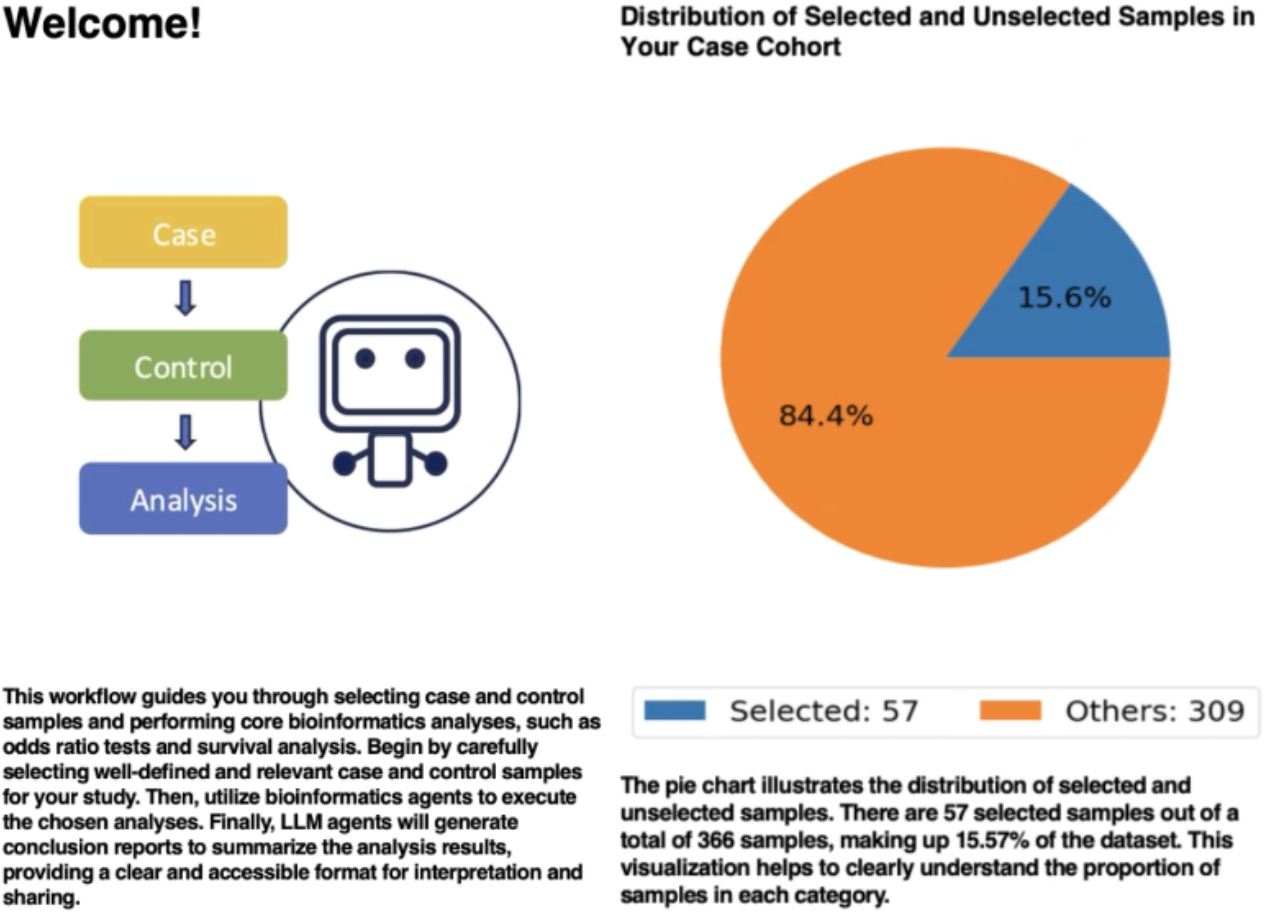
AI-HOPE Case-Control Study Comparing TP53 Mutations in Early-Stage (I/II) and Late-Stage (III/IV) CRC Patients. The figure consists of two panels: a “Welcome” panel on the left and a pie chart on the right. The left panel introduces the AI-HOPE workflow, guiding users through the process of selecting case and control samples, performing bioinformatics analyses such as odds ratios and survival analysis, and generating actionable insights. It emphasizes the role of LLM agents in automating the process and providing clear, interpretable results. The right panel features a pie chart illustrating the sample distribution from a case-control study conducted by AI-HOPE using colorectal cancer (CRC) data from The Cancer Genome Atlas (TCGA). Out of 365 total samples analyzed, 57 samples (15.6%) were classified into the early-stage (I/II) group, while the remaining 309 samples (84.4%) represent unselected samples or other stages.

In another demonstration, AI-HOPE compared FOLFOX-treated CRC patients with or without RAS mutations. The system detected a significant difference in progression-free survival, identifying that KRAS mutations were predictive of a poorer response to FOLFOX treatment and an increased risk of recurrence. These findings were consistent with those reported by Nicolantonio et al. (2021), underscoring AI-HOPE’s ability to independently uncover clinically relevant insights (Fig. 3).

**Figure 3:**
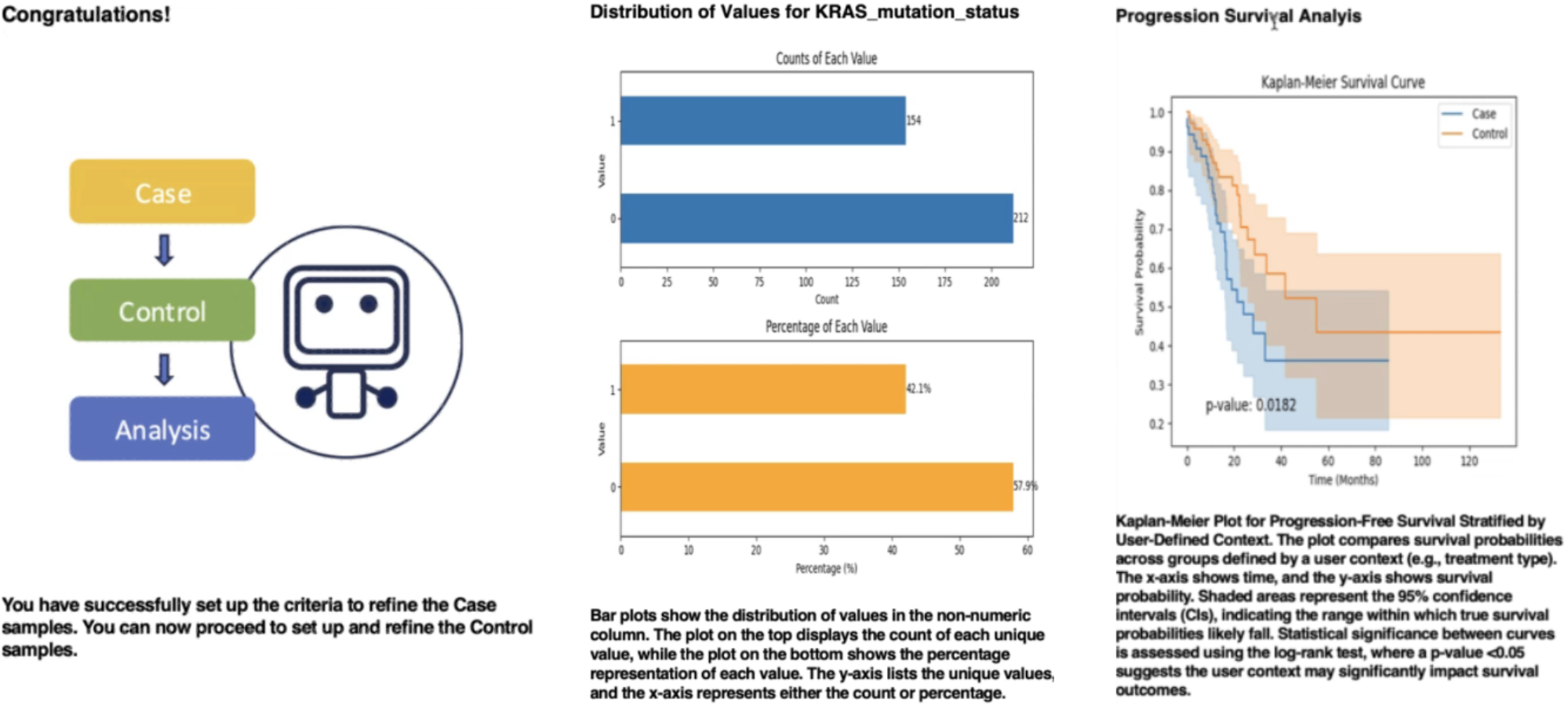
AI-HOPE Analysis of FOLFOX-Treated CRC Patients with and without RAS Mutations. This figure is composed of three panels—left, center, and right— highlighting key stages in AI-HOPE’s analysis of progression-free survival among FOLFOX-treated colorectal cancer (CRC) patients with or without RAS mutations. The left panel provides an overview of the AI-HOPE workflow, indicating that the criteria for refining case samples have been successfully set. It emphasizes the readiness to proceed with further analysis, such as the definition and refinement of control groups and subsequent bioinformatics analyses. The central panel presents bar charts illustrating the distribution of KRAS mutation statuses within the analyzed cohort. The top chart shows the count of samples for each mutation status, while the bottom chart displays the corresponding percentage distribution. This visualization provides a clear summary of KRAS mutation frequencies, forming the foundation for subsequent survival analysis. The right panel features a Kaplan-Meier survival curve, stratified by KRAS mutation status, to evaluate progression-free survival among FOLFOX-treated patients. The plot demonstrates that patients with KRAS mutations had significantly poorer progression-free survival compared to those without KRAS mutations, as indicated by the p-value and confidence intervals.

Overall, these examples highlight AI-HOPE’s potential to autonomously perform complex analyses and generate meaningful interpretations, enabling researchers to efficiently derive conclusions from clinical and genomic data without requiring technical expertise.

## Discussion

The Artificial Intelligence agent for High-Optimization and Precision mEdicine (AI-HOPE) represents a transformative step in precision medicine research by combining advanced machine learning capabilities with user-friendly natural language processing. This study demonstrates AI-HOPE’s ability to streamline clinical and genomic data analyses, significantly lowering the barrier to entry for researchers without programming expertise. By autonomously executing complex analytical workflows, including subset selection, prevalence analysis, and survival analysis, AI-HOPE exemplifies the potential of artificial intelligence to enhance efficiency and precision in clinical research.

A key strength of AI-HOPE lies in its capacity to conduct user-defined case-control studies, as demonstrated in our CRC examples. These analyses highlighted the enrichment of TP53 mutations in late-stage CRC and revealed a significant association between KRAS mutations and poorer progression-free survival in FOLFOX-treated patients. Not only were these findings consistent with established literature, but they also underscored the system’s ability to independently identify and validate clinically meaningful insights.

The versatility of AI-HOPE extends beyond CRC research, offering applications across a wide spectrum of cancer types and potentially other diseases. By allowing flexible stratification based on clinical and molecular attributes, AI-HOPE can empower researchers to identify biomarkers, evaluate treatment outcomes, and explore disease mechanisms more comprehensively. The ability to integrate and analyze diverse datasets, such as those from TCGA and cBioPortal, further enhances the tool’s utility across research domains.

While AI-HOPE offers remarkable promise, there are areas for further development. Future iterations could incorporate more advanced natural language processing to handle increasingly complex queries and integrate larger, real-time datasets.

Additionally, benchmarking AI-HOPE against traditional bioinformatics pipelines will provide a clearer understanding of its efficiency and accuracy. Expanding its functionality to include multi-omics data integration and prediction modeling could further enhance its utility in precision medicine research.

Overall, AI-HOPE demonstrates the potential of conversational AI to revolutionize clinical and genomic research. By simplifying data analysis workflows and providing actionable insights, AI-HOPE serves as a valuable tool to bridge the gap between data complexity and research needs, ultimately accelerating discoveries in precision medicine research.

## Conclusion

The Artificial Intelligence agent for High-Optimization and Precision mEdicine (AI-HOPE) represents a significant advancement in precision medicine research, addressing critical challenges in integrating clinical and genomic data. By harnessing the power of LLMs, AI-HOPE offers a user-friendly, conversational interface that empowers researchers to perform sophisticated analyses without requiring programming expertise. Its demonstrated success in case-control studies, such as identifying TP53 mutations in CRC and assessing KRAS mutations in treatment outcomes, underscores its potential to generate actionable insights that align with established scientific findings. Beyond cancer studies, AI-HOPE’s versatile and adaptable design positions it as a valuable tool for a broad spectrum of biomedical research. Its robust analytical capabilities enable the integration of omics data, clinical metadata, and treatment outcomes, providing a scalable framework that addresses critical needs in translational research. This adaptability makes AI-HOPE suitable for exploring biomarkers, understanding disease mechanisms, and evaluating therapeutic responses across various complex diseases. Looking ahead, AI-HOPE has the potential to evolve further by incorporating advanced natural language processing, real-time data integration, and multi-omics analysis capabilities. These enhancements will enhance its utility, solidifying its role as a transformative resource in precision medicine. By bridging the gap between data complexity and research needs, AI-HOPE accelerates discoveries and offers a powerful platform to improve patient outcomes across diverse fields of biomedical science.

## Data Availability Statement

All data used in the present study is publicly available at https://www.cbioportal.org/ (accessed on 8 September 2024), https://genie.cbioportal.org (accessed on 8 September 2024) and https://xena.ucsc.edu (accessed on 8 September 2024)]. Additional data can be provided upon reasonable request to the authors.

